# Low perfusion and missed diagnosis of hypoxemia by pulse oximetry in darkly pigmented skin: A prospective study

**DOI:** 10.1101/2022.10.19.22281282

**Authors:** M Koa Gudelunas, Michael Lipnick, Carolyn Hendrickson, Sky Vanderburg, Bunmi Okunlola, Isabella Auchus, John R. Feiner, Philip E. Bickler

## Abstract

**Importance:** Retrospective clinical trials of pulse oximeter accuracy report more frequent missed diagnoses of hypoxemia in hospitalized Black patients than White patients, differences that may contribute to racial disparities in health and health care. Retrospective studies have limitations including mistiming of blood samples and oximeter readings, inconsistent use of functional versus fractional saturation, and self-reported race used as a surrogate for skin color. Understanding the cause of biased readings by pulse oximetry in patients with darkly pigmented skin is high priority given the essential nature of pulse oximetry.

**Objective:** To prospectively measure the contributions of skin pigmentation, perfusion index, sex, and age on pulse oximeter errors.

**Design:** We studied two pulse oximeters (Nellcor N-595™ and Masimo Radical 7™) in prevalent use in North America, Europe, and Asia-Pacific regions. We analyzed 9,763 matched pulse oximeter readings (SpO2) and arterial oxygen saturation (hemoximetry SaO2) during stable hypoxemia (SaO2 68-100%). Perfusion index (PI) was measured as percent infrared light modulation by the pulse detected by the pulse oximeter probe, with low perfusion categorized as PI <1%.

**Setting:** Clinical research laboratory

**Participants:** 146 healthy subjects, including 25 with light skin (Fitzpatrick class I-II), 78 with medium (class III-IV), and 43 with dark (class V-VI) skin.

**Exposures:** Controlled hypoxemia

**Main Outcomes:** Pulse oximeter bias (difference between SaO2 and SpO2) by skin pigment category in a multivariable mixed-effects model incorporating repeated-measures and different levels of SaO2 and perfusion.

**Results:** Skin pigment, perfusion index and degree of hypoxemia significantly contributed to errors (bias) in both pulse oximeters. The combined frequency of missed diagnosis of hypoxemia (pulse oximeter readings 92-96% when arterial oxygen saturation was <88%) in low perfusion conditions was 1.1% for light, 8.2% for medium and 21.1% for dark skin.

**Conclusions and Relevance:** Low peripheral perfusion combined with darker skin pigmentation leads to clinically significant high-reading pulse oximeter errors and missed diagnoses of hypoxemia. Darkly pigmented skin and low perfusion states are likely the cause of racial differences in pulse oximeter performance in retrospective studies. Both skin pigmentation and low perfusion should be accounted for in regulatory standards for pulse oximeters.

**Key Points:** *Question:* Laboratory validation of pulse oximeter performance has found errors in Black subjects that are smaller than those from recent reports of hospitalized Black patients. We test the hypothesis that low perfusion amplifies pulse oximeter error in subjects with dark skin during hypoxemia.

*Findings:* Dark skin pigmentation combined with low perfusion produces large errors in pulse oximeter readings in healthy subjects during hypoxemia, sufficient to miss a diagnosis of hypoxemia in about 20% of readings.

*Meaning:* Accuracy of pulse oximeters in the diagnosis of hypoxemia is impaired by a combination of low perfusion and dark skin pigmentation. Low perfusion should be accounted for in future testing and regulatory guidelines for pulse oximeters to improve performance and minimize bias in patients with dark skin pigment.

## Introduction

Pulse oximeters are indispensable diagnostic tools worldwide, and small errors in performance could lead to significant impacts on health and healthcare. Pulse oximeters cleared by the United States Food and Drug Administration (FDA) must read within 3% of arterial saturation (functional saturation, SaO2), but oximeter performance is degraded by the presence of numerous factors including skin melanin, dyshemoglobins, anemia, motion, and low perfusion^1,2^. Validation studies of pulse oximeter performance for International Organization for Standardization (ISO) or FDA clearance exclusively involve healthy young adult subjects with minimal co-morbidities that could interfere with device performance^3,4^. Since documentation of missed hypoxemia diagnosis due to melanin was documented by Bickler *et al*. and Feiner *et al*. in 2005 and 2007^5,6^, the FDA has required inclusion of subjects with darkly pigmented skin in validation studies^7^. However, there is growing concern and evidence to suggest that current healthy human, laboratory-based study validation protocols may not adequately predict performance in real-world clinical settings^8^. Recent retrospective studies report missed diagnoses of hypoxemia in patients with darkly pigmented skin at twice the rate compared to White patients^9-12^. These findings have prompted inquiries by multiple regulatory bodies worldwide, including letters from Congress to the FDA and a new FDA communication^13,14^. Errors in pulse oximetry in hospitalized patients may lead to worse clinical outcomes and a perpetuation of disparities in health and healthcare^15,16^. Because the magnitude and cause of these errors in real-world clinical settings remain uncertain, the path to resolution is unclear.

Previous studies in our laboratory found that low finger perfusion, measured by modulation of infrared light transmission through a finger, is associated with increased pulse oximeter errors^17^. The authors have also observed that darkly pigmented skin is associated with more frequent pulse oximeter errors under low perfusion conditions, both in healthy human subjects and in patients who are critically ill, in shock, on vasoconstrictive medications or those with vascular disease. The purpose of this study was to test the hypothesis that pulse oximeter SpO2 overestimates SaO2 more frequently in the presence of increased skin melanin *and* low perfusion than in the presence of either condition alone. We studied two pulse oximeters that represent a significant share of pulse oximeters used in North American hospitals in 2022^18^. This study was specifically designed to overcome important confounding factors present in retrospective trials, including mistiming of blood samples and oximeter readings, inconsistent use of functional versus fractional saturation, and self-reported race used as a surrogate for skin color^8^.

## Methods

We used data from 146 consecutive, healthy, nonsmoking volunteer participants in pulse oximeter performance studies at the UCSF Hypoxia Research Laboratory in 2020 and 2021. Demographic information on the study population is presented in Table 1. Written informed consent was obtained from all subjects under IRB approval 10-00437 from UCSF. As the study did not assign subjects to different treatments for assessment of health outcomes, it does not meet the definition of a clinical trial for registration at clinicaltrials.gov.

**Table 1.** Demographic information and sample distribution.

| n | 146 | Samples |
| --- | --- | --- |
| Age (years) | 27.7 ± 5.6 | 9,763 |
| Gender |  |  |
| Female | 77 (52.7%) | 4,316 |
| Male | 69 (47.3%) | 5,447 |
| Ethnicity/Race |  |  |
| Asian | 47 (32.2%) | 2,948 |
| Black | 23 (15.8%) | 1,469 |
| Caucasian | 42 (28.8%) | 3,197 |
| Hispanic | 20 (13.7%) | 1,125 |
| Multiethnic | 14 (9.6%) | 1,024 |
| Fitzpatrick scale |  |  |
| I | 3 (2.1%) | 175 |
| II | 22 (15.1%) | 1,599 |
| III | 41 (28.1%) | 2,172 |
| IV | 37 (25.3%) | 2,961 |
| V | 23 (15.8%) | 1,683 |
| VI | 20 (13.7%) | 1,173 |

Data are mean ± SD, n or n (%)
Ethnicity, self-reported; samples include all arterial blood gas samples.

### Skin pigment assessment and Demographics

Each subject’s skin pigmentation was classified by the Fitzpatrick scale, commonly used in studies of pulse oximetry performance following both FDA and ISO standards^19-21^. Race, ethnicity and sex were self-reported.

### Perfusion index

The perfusion index (PI) from each oximeter probe was recorded continuously. The Nellcor PI was divided by 10 to make the values numerically comparable to Masimo. Since no standard exists to define low perfusion conditions for pulse oximetry, a sensitivity analysis comparing bias and root mean square error (Arms) at various levels of PI was performed. Mean bias in oximeter readings increased approximately 5-fold in grouping of readings with PI >2 versus PI<0.5 %. Scatterplots revealed that the relationship of bias to PI becomes very small at high values (PI≥ 2%) in a non-linear fashion. For groups of data with PI values of 1.0-1.5 %, 0.5-1.0 % and <0.5%, the P value of the relationship to mean bias or median absolute bias was <0.00001. Accordingly, low perfusion was defined for the purpose of this study as PI< 1.0%.

### Hypoxemia protocol

Participants were in a 30-degree head up semi-recumbent position for the study. After local anesthesia, a 22-ga catheter was placed in a radial artery. To create steady-state hypoxemia, subjects breathed controlled air-nitrogen-CO2 mixtures via a mouthpiece and partial re-breathing system to achieve multiple stable plateaus of arterial oxygenation and a relatively constant end-tidal CO2 per ISO and FDA guidelines^19,20^. A more detailed description of testing the accuracy of pulse oximetry devices in the laboratory setting has been published^3,4^. As is common practice in laboratory-based studies, warming pads on the hands and/or forearms were used on some subjects to improve perfusion, but no particular level of perfusion was sought, and subjects with low perfusion were not removed from the study. Blood samples for SaO2 measurement (Radiometer ABL90 Flex, Copenhagen, Denmark) were collected through the radial artery catheter.

A Masimo Radical 7 (Non-touch screen model, Masimo Inc., Irvine, CA) with a DCI clip-type adult reusable finger sensor or ear clip sensor, and Nellcor N-595 (Medtronic Inc, Minneapolis, MN) with a reusable finger clip sensor (Nellcor DS-100A) were placed on one of the middle three fingers or ear. Probes were re-positioned as necessary to ensure proper placement throughout the study. No subjects wore nail polish. Data from the pulse oximeters were recorded at 2 Hz from the instruments’ serial output ports using a computer running LabVIEW 15.0. (National Instruments, Austin, Tx). Data from the Masimo were transmitted at 1 Hz, while the data from the Nellcor were transmitted at 0.5 Hz. Recorded data included SpO2, heart rate and perfusion index from each device.

Stable SaO_2_ plateaus between 60% and 100% were targeted by the study physician who adjusted the inspired gas mixture. At each level, two arterial blood samples were collected, approximately 30 seconds apart, each during steady-state conditions. Some (233 of 9763) SpO_2_ values were eliminated for obvious oximeter signal dropout, or failure to reach an appropriate stable plateau. Only samples with stable SpO_2_ readings (i.e., fluctuation <2% per minute) were included in the analysis. No subjects had MetHb or COHb outside the normal range as measured by the hemoximeter (Radiometer ABL90 Flex, Copenhagen, Denmark).

### Statistics

Bias (or error) was computed as SpO_2_ minus the corresponding arterial blood SaO_2_. Bias assessment parameters included the mean bias (“accuracy”), standard deviation of the bias (“precision”), and root mean square error (A_rms_)—an overall performance measure used by the FDA. A_rms_ was calculated as the square root of the mean difference between SpO_2_ and SaO_2_, squared. The 95% confidence limits of A_rms_ were determined using bootstrapping (random resampling with replacement) with 50,000 repetitions. Prior sensitivity analyses showed that this was sufficient, as the results did not change at the reported level of significance. The 95% limits of agreement (LOA) were calculated as 1.96 •SD according to Bland and Altman agreement analysis with adjustments for multiple measurements for each individual according to the “Method Where the True Value Varies”^22^. The 95% confidence limits for the LOA were also determined using bootstrapping.

The primary analysis was a multivariable mixed-effects model of bias that accounts for repeated-measures changes in individuals at different SaO_2_ levels and different perfusion index levels, with fixed effects for gender and skin classification. Three skin pigment categories (light medium and dark) were used in the model for clarity, but results were confirmed with the six Fitzpatrick skin classifications. A three-level perfusion index scale was used: PI<1%, PI ≥1% and <2%, and PI≥2%. A complete model with all terms, and including all possible interactions of skin pigmentation, PI and SaO_2_, was performed to confirm the results. Analysis was performed in both MatLab (The Mathworks, Inc., Boston, MA) and Stata 17.0 (Statacorp., College Station, TX).

Given the possibility of confounding, possible interactions between the variables in the model were also analyzed. Interactions between PI and both sex and three skin categories were assessed using the Wilcoxon rank sum and Kruskal-Wallis tests, respectively. Age and gender were not significantly related to bias, but PI differed between women (median [IQR]: 2.0 [0.8-3.5]) and men (4.5 [2.0-7.4]), *P*<0.0001. Perfusion index did not vary between light, medium and dark skin categories, *P*=0.85. The distribution of skin pigmentation categories did not differ between men and women, *P*=0.41

Receiver operator characteristic (ROC) curves were constructed for the definition of hypoxemia (SaO_2_ < 88%) at various SpO_2_ thresholds for the Masimo and Nellcor pulse oximeters. “Missed hypoxemia” was defined as the percentage of pulse oximeter readings with SpO_2_ at or above cutoff values of 92%, 94% and 96% with corresponding SaO_2_ < 88%.

## Results

### Study population and sample size

We used data from 146 consecutive, healthy, nonsmoking volunteer participants in pulse oximeter performance studies at the UCSF Hypoxia Research Laboratory in 2020 and 2021. Demographic information on the study population is presented in Table 1. Forty-three subjects classified as Fitzpatrick skin type V or VI (Dark), 78 as Fitzpatrick III or IV (Medium) and 25 as Fitzpatrick I or II (Light). A total of 9,763 arterial blood gas samples were analyzed with 1,774 from Fitzpatrick I-II, 5,133 from Fitzpatrick III-IV, and 2,856 samples from Fitzpatrick V-VI volunteers (Table 1). Each subject participated in an average (SD) of 2.8 ± 3.0 studies (median [IQR]: 1 [1-3]; range 1-16) studies with 65 subjects who had data from ≥1 study.

### Primary Outcomes

A multivariable analysis accounting for repeated measures revealed that skin pigment, perfusion index and degree of hypoxemia significantly contributed to the amount of error (bias) in both pulse oximeters (Table 2) as significant interaction was found between these three variables. Supplemental Tables (eTables 1, 2 and 3) summarize bias statistics for each of the variables separately. For both pulse oximeters, mean bias and Arms increased with skin pigmentation class, and increased with decreasing perfusion. Since gender was highly correlated to PI, the model was also run separately for men and women with the same result (data not shown).

**Table 2.**
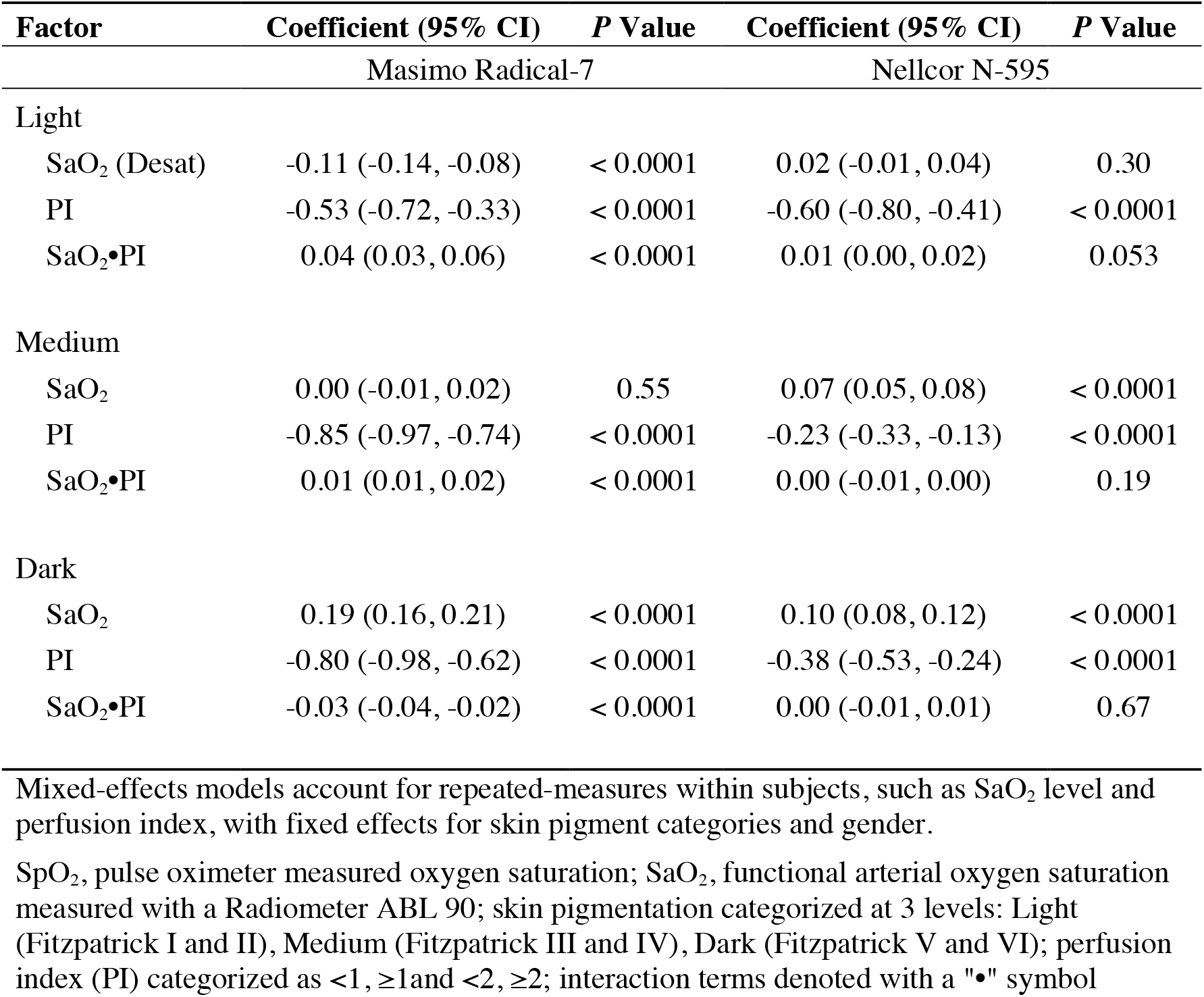
Contributions of saturation range, skin pigment, perfusion level and interactions to pulse oximeter bias. Mixed-effects multivariable models

| Factor | Masimo Radical-7 |  | Nellcor N-595 |  |
| --- | --- | --- | --- | --- |
|  | Coefficient (95% CI) | P Value | Coefficient (95% CI) | P Value |
| Light |  |  |  |  |
| SaO<sub>2</sub> (Desat) | -0.11 (-0.14, -0.08) | < 0.0001 | 0.02 (-0.01, 0.04) | 0.30 |
| PI | -0.53 (-0.72, -0.33) | < 0.0001 | -0.60 (-0.80, -0.41) | < 0.0001 |
| SaO<sub>2</sub>•PI | 0.04 (0.03, 0.06) | < 0.0001 | 0.01 (0.00, 0.02) | 0.053 |
| Medium |  |  |  |  |
| SaO<sub>2</sub> | 0.00 (-0.01, 0.02) | 0.55 | 0.07 (0.05, 0.08) | < 0.0001 |
| PI | -0.85 (-0.97, -0.74) | < 0.0001 | -0.23 (-0.33, -0.13) | < 0.0001 |
| SaO<sub>2</sub>•PI | 0.01 (0.01, 0.02) | < 0.0001 | 0.00 (-0.01, 0.00) | 0.19 |
| Dark |  |  |  |  |
| SaO<sub>2</sub> | 0.19 (0.16, 0.21) | < 0.0001 | 0.10 (0.08, 0.12) | < 0.0001 |
| PI | -0.80 (-0.98, -0.62) | < 0.0001 | -0.38 (-0.53, -0.24) | < 0.0001 |
| SaO<sub>2</sub>•PI | -0.03 (-0.04, -0.02) | < 0.0001 | 0.00 (-0.01, 0.01) | 0.67 |

Mixed-effects models account for repeated-measures within subjects, such as SaO<sub>2</sub> level and perfusion index, with fixed effects for skin pigment categories and gender.
SpO<sub>2</sub>, pulse oximeter measured oxygen saturation; SaO<sub>2</sub>, functional arterial oxygen saturation measured with a Radiometer ABL 90; skin pigmentation categorized at 3 levels: Light (Fitzpatrick I and II), Medium (Fitzpatrick III and IV), Dark (Fitzpatrick V and VI); perfusion index (PI) categorized as <1, ≥1 and <2, ≥2; interaction terms denoted with a "•" symbol

The interaction between hypoxemia and PI in darkly pigmented subjects shows that positive bias increases synergistically during hypoxemia with lower perfusion, an effect seen to a smaller extent in medium and lightly pigmented subjects (Fig. 1). The shift in distribution of pulse oximeter readings towards larger errors during low perfusion differed between subjects with light, intermediate and dark skin (Fig. 2).

**Figure 1.**
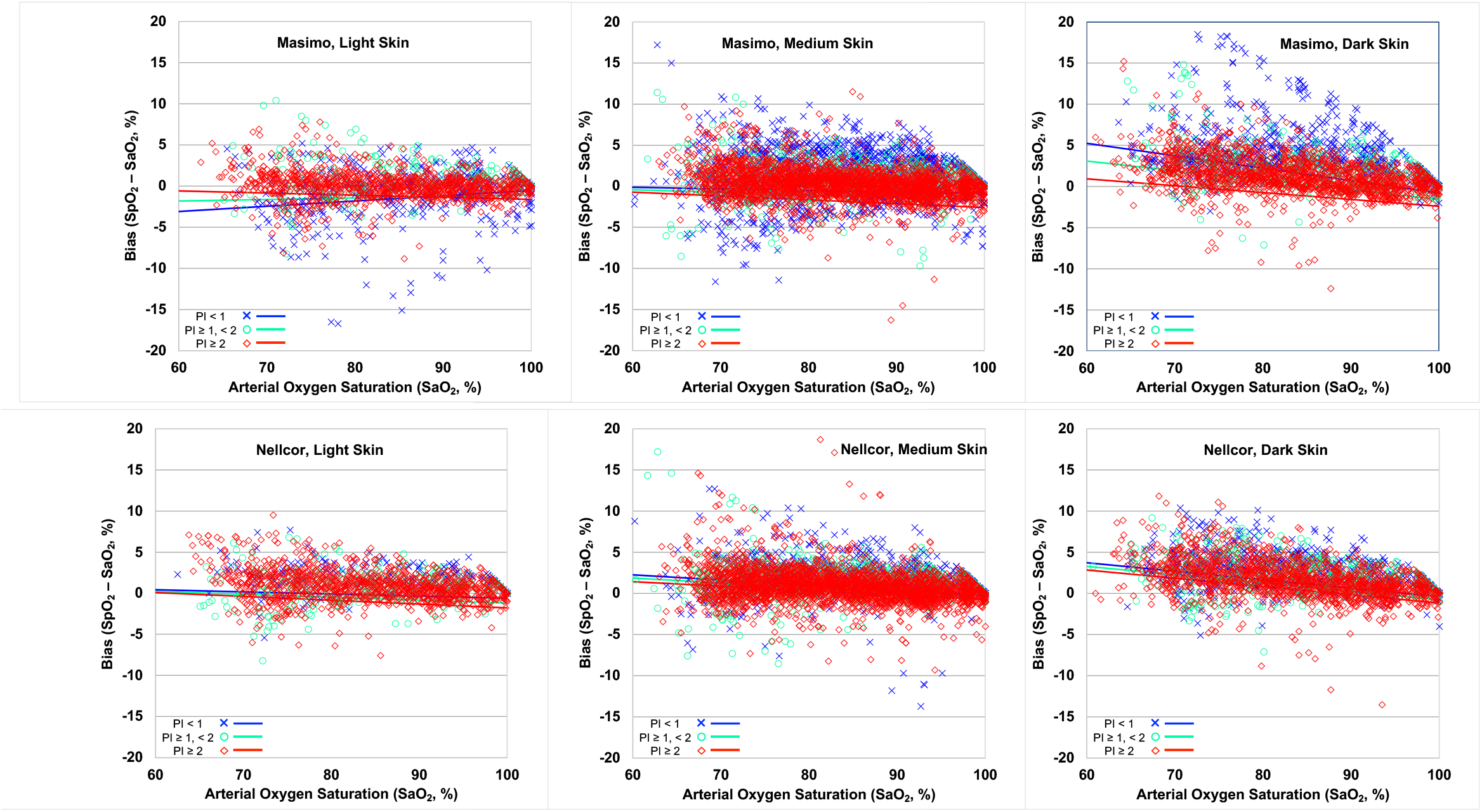
Pulse oximeter bias (SpO_2_ -SaO_2_) vs. SaO_2_ (measured arterial blood oxygen saturation) in light (Fitzpatrick I and II), medium (Fitzpatrick III and IV) and darkly (Fitzpatrick V and VI) pigmented human subjects. Regression lines at different level of perfusion index (PI) are shown separately (Blue: PI<1%; Purple: PI≥1% and <2%; Pink: PI≥2%). Values for the slopes with 95% confidence intervals are shown on the graphs. Data were analyzed by a mixed-effects multivariable model accounting for repeated measures.

**Figure 2.**
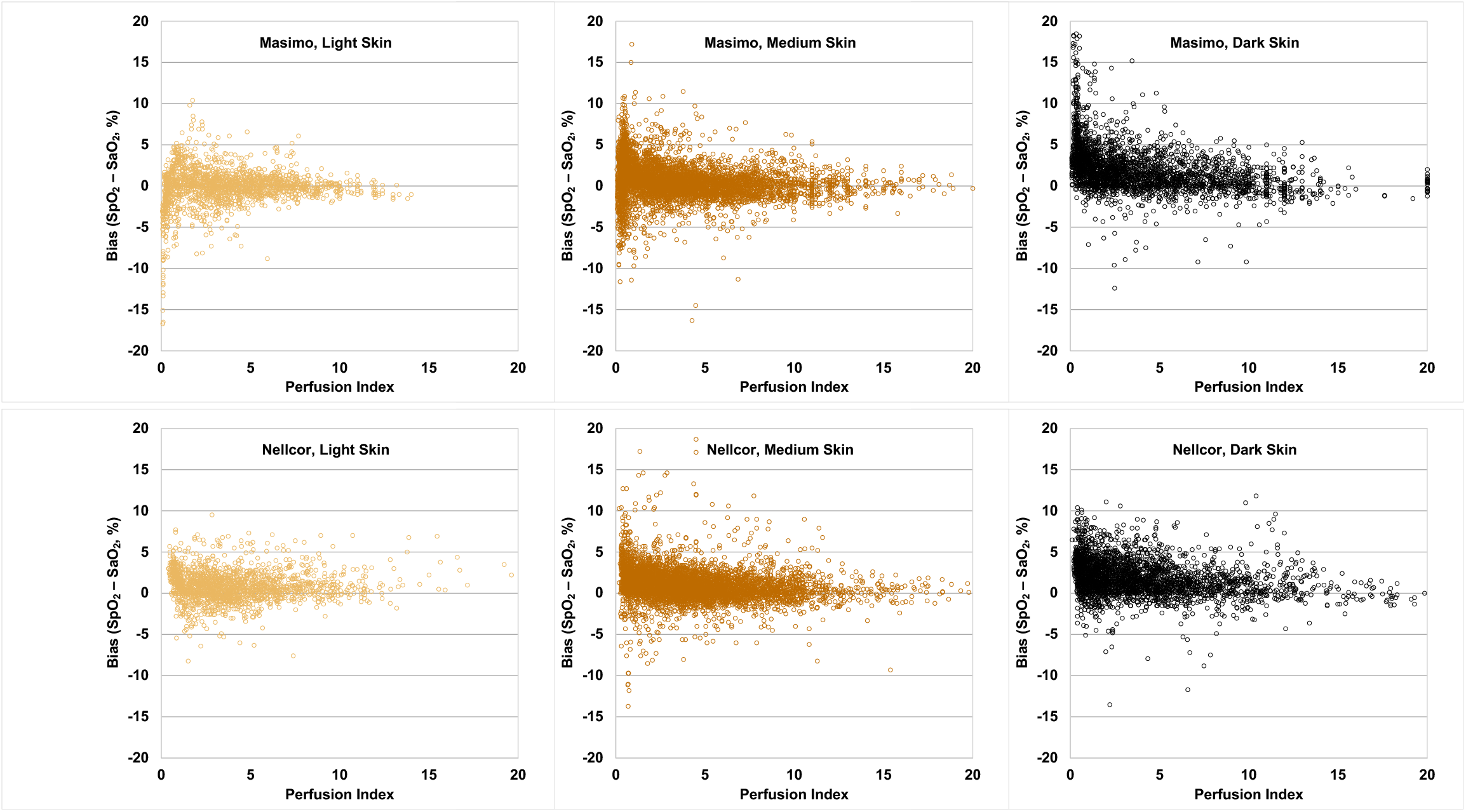
*Upper panels.* Relationship of perfusion index (PI, % modulation in infrared light through the finger) with the Masimo Radical Probe and pulse oximeter bias in subjects by skin pigment class. *Lower panels*. Bias (SaO_2_ - SpO_2_) with mean ±SD of Nellcor N-595 and Masimo Radical pulse oximeter readings segregated by perfusion index (PI<1%, PI≥1% and <2%, or ≥2%) and Fitzgerald skin pigment class. * P<0.01, ** P<0.001, *** P<0.0001 compared to bias in Fitzgerald class I-II subjects with normal perfusion.

### Errors in pulse oximetry and missed diagnosis of hypoxemia

Pulse oximeter errors near the threshold for the diagnosis of hypoxemia (SaO_2_ 88-90%) have special clinical importance. In Fig. 3, we compare pulse oximeter readings and corresponding SaO_2_, identifying values for which the pulse oximeter read 92% or greater when SaO_2_ was <88%, representing a missed hypoxemia diagnosis. Missed hypoxemia was more frequent with darker skin and lower perfusion for both oximeters tested. For subjects with darkly pigmented skin and low perfusion, missed hypoxemia for the SpO_2_ range of 92-96%was seen in 30.2% of the readings from the Masimo pulse oximeter and 7.9% of the readings from the Nellcor. The same trend was seen when different thresholds for errors were analyzed. The frequency of missed hypoxemia for SpO_2_ thresholds of ≥92%, ≥94%, ≥96% and 92-96% are summarized in eTable 4.

**Figure 3.**
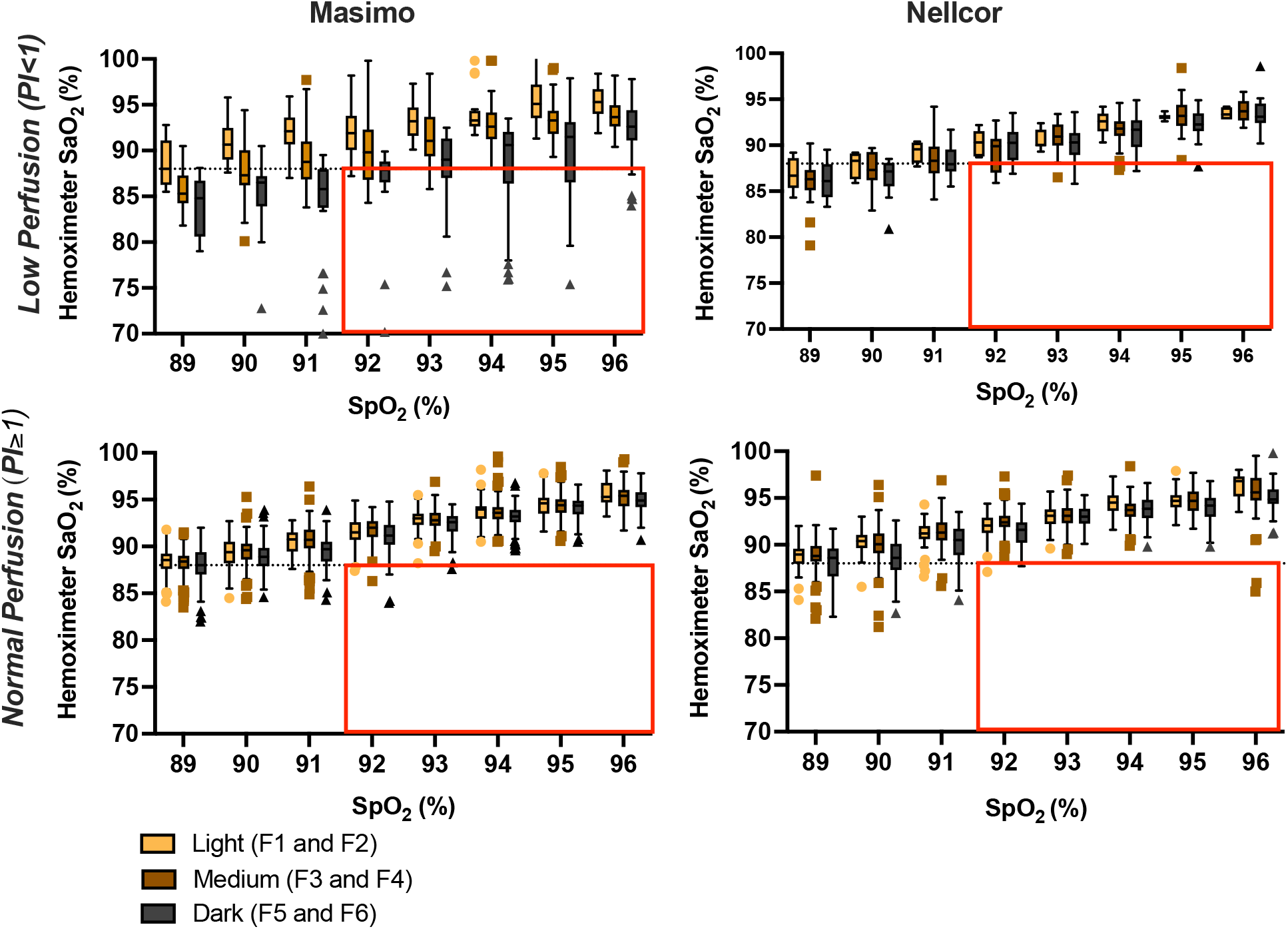
Analysis of “missed hypoxemia”, defined as a hemoximeter SaO_2_ of <88% with a corresponding pulse oximeter reading of 92% or greater. Data are separated by skin pigment classification and perfusion level, with low perfusion defined as PI<1, see text. The dashed lines show an arterial oxygen saturation of 88%. The red rectangular area delimits data pairs defined by missed hypoxemia. In the box plots, the horizontal line within each box represents the median, the top and bottom of the box represent the upper and lower limits of the interquartile range, and the whiskers represent 1.5 times the interquartile range. Outliers outside these ranges are indicated by single data points. Rates of missed diagnosis for normal and low perfusion by skin class are in supplemental Table 4.

The area under the receiver operating characteristic curve (ROC) for a diagnosis of hypoxemia was lowest for subjects with darkly pigmented skin and low perfusion, 0.96 (95% CI: 0.95-0.98). Sensitivity cutoff values were shifted to higher perfusion values in subjects with dark skin. For example, to achieve 95% sensitivity for SaO_2_ < 88% for the Masimo in darkly pigmented subjects with PI<1%, a cutoff of ≤95% would be needed, whereas this cutoff would correspond to 99% sensitivity for SaO2 < 88% in darkly pigmented subjects with PI ≥2% (data not shown).

## Discussion

In this prospective study of healthy volunteers, we have found that low perfusion interacts with both medium and dark skin pigment to substantially increase pulse oximeter bias. For subjects with dark skin and low perfusion, these errors lead to missed hypoxemia diagnoses at frequencies comparable to or higher than those reported for Black hospitalized patients in retrospective clinical studies^9-12^. Because critical illness is commonly associated with low peripheral perfusion, this finding may be of significant clinical importance. A prospective clinical trial of pulse oximeter bias in relation to skin pigment, perfusion level and degree of hypoxemia in hospitalized patients is underway to confirm this association ^23^.

We have identified a mechanism by which pulse oximeter errors seen in laboratory conditions in humans may be amplified in real world clinical environments. Our findings therefore suggest that the misdiagnosis of hypoxemia in hospitalized patients in previous retrospective studies cannot simply be dismissed because of potential study design limitations including uncertain synchronization of blood draws and stable pulse oximeter readings, interfering compounds in the blood, etc^8^. If such errors confounded retrospective studies, an even distribution of positive and negative bias might be expected. However, this study, previous laboratory studies ^5,6,8^ and the retrospective hospital studies^9-11,15,16,24^ find positive pulse oximeter bias in subjects with darker skin. In this laboratory-based study involving a large number of samples in the 70-80% saturation range, we confirm that the positive bias becomes larger at low saturation^5,6^.

### Pulse oximeter bias and missed hypoxemia diagnosis

Missed diagnosis of hypoxemia occurred at concerning rates both for subjects with not only dark skin pigment but also medium skin pigment, which is a new finding. The combination of dark skin and low perfusion causes errors in pulse oximeter readings large enough to miss the diagnosis of hypoxemia in 8-30% of readings when SaO_2_ is <88% and the pulse oximeter reads 92-96% (Supplemental Table 4). In darkly pigmented subjects across all ranges of perfusion, the missed diagnosis rate was about 3-9%, and for subjects with medium skin pigmentation, the missed diagnosis rate was about 1-2%. Subjects with medium skin pigment in our study self-identified as Asian, Hispanic, or Multiethnic. Our findings remain robust when different ranges of hypoxemia are used as the threshold for a finding of missed diagnosis (Supplemental Table 4).

### Low Perfusion and clinically significant pulse oximeter errors

Our thresholds for low perfusion were selected based on a sensitivity analysis of the relationship of PI to level of pulse oximeter bias. In our clinical experience in the ICU and perioperative settings, low peripheral perfusion is commonly encountered due to underlying disease as well as lack of routine warming of patient hands during oximetry monitoring. Poor peripheral perfusion is common in the elderly and in patients with sepsis, peripheral vascular disease, hypertension, diabetes, and other conditions^25^. The incidence of low perfusion readings was not reported in any of the recent clinical studies concerning pulse oximeter errors in hospitalized patients.

An analysis of pulse oximeter errors in surgical patients by Burnett *et al*. is relevant to the issue of how perfusion level affects pulse oximeter bias^10^. Due to the vasodilatory actions of anesthetics and use of forced air warming in the operating room, peripheral perfusion in surgical patients is expected to be greater than in other hospitalized patients. Therefore, we are not surprised that the incidence reported by Burnett of missed diagnosis of hypoxemia in black patients is about half that in non-surgical patient cohorts reported by Sjoding, Valbuena, and others^9-12^.

Our results suggest that pulse oximeter errors in darkly pigmented humans are not exclusively related to skin pigmentation interference with light signals. Unless pulse oximeter electronic and optical systems adequately account for decreased pulse signals and other physiologic changes during low perfusion, a simple colorimetry-based calibration-correction for pulse oximeter bias is unlikely to work. Not only are the physio-optic challenges complex, they are dynamic. For example, with good perfusion, errors in darkly pigmented subjects are nearly comparable to those in lightly pigmented subjects (Fig. 1).

### Regulatory implications

Significant pulse oximeter bias occurring in subjects with medium and dark skin pigment and low perfusion was obscured when overall performance (i.e., all subjects) of the pulse oximeters was analyzed (eFigure 1). Both instruments performed within the 3% specification for FDA clearance. Currently, the FDA requires pulse oximeters to be tested in a minimum of 2 subjects or 15% with dark skin in a population of 10 subjects, whichever is larger. Based on this study, we propose that pulse oximeters should be tested in subjects over a range of skin pigmentation classifications and with low perfusion, specifically with PI <1%. Such testing could take place in real world clinical trials either pre- or post-market but should also take place in pre-market laboratory-based healthy volunteer studies. By including low perfusion enrollment targets in laboratory-based validation studies, these tests are more likely to predict real world performance, decrease bias and improve equity.

### Study limitations: laboratory vs clinical settings

This study has several limitations. First, this study enrolled young healthy subjects, and thus cannot be equated with data from real world clinical settings. However, the physiological control with stable levels of hypoxemia made possible by this study design produced better synchronization of stable SpO_2_ and measured SaO_2_ than is possible in retrospective clinical studies. Moreover, multiple data points for different individuals under different conditions allowed for an extremely robust analysis of the impacts of perfusion on bias. Of note, the distribution of perfusion in hypoxemic hospitalized patients has not been established. Second, this study utilized Fitzpatrick scale for skin color assessment. Due to the many previously identified limitations of this method, we have shifted to other quantitative skin pigment assessments for future studies^8,26^. Finally, we studied two pulse oximeter models widely used in the clinical setting; however, the manufacturers of these pulse oximeters have other models— including newer models—which may have different performance characteristics. Consequently, generalization of these data to other pulse oximeter manufacturers and models is uncertain.

## Conclusions

Low perfusion is associated with misdiagnosis of arterial hypoxemia by pulse oximetry in healthy subjects with dark skin pigmentation in controlled laboratory conditions. The frequency of misdiagnosis of clinically significant hypoxemia in our laboratory study was similar to that observed in Black patients in recent retrospective clinical studies. Because low perfusion is common in critically ill patients, clinicians that rely on pulse oximetry alone to diagnose and treat hypoxemia need to recognize that missed diagnosis increases greatly at low perfusion, particularly in patients with medium or darkly pigmented skin. Pulse oximeter regulatory guidance must be updated to better account for oximeter performance not only in patients with dark skin pigment but also during low perfusion states.

## Data Availability

All data produced in the present study are available upon reasonable request to the authors

## Summary of Supplemental Content

**eTable 1.**
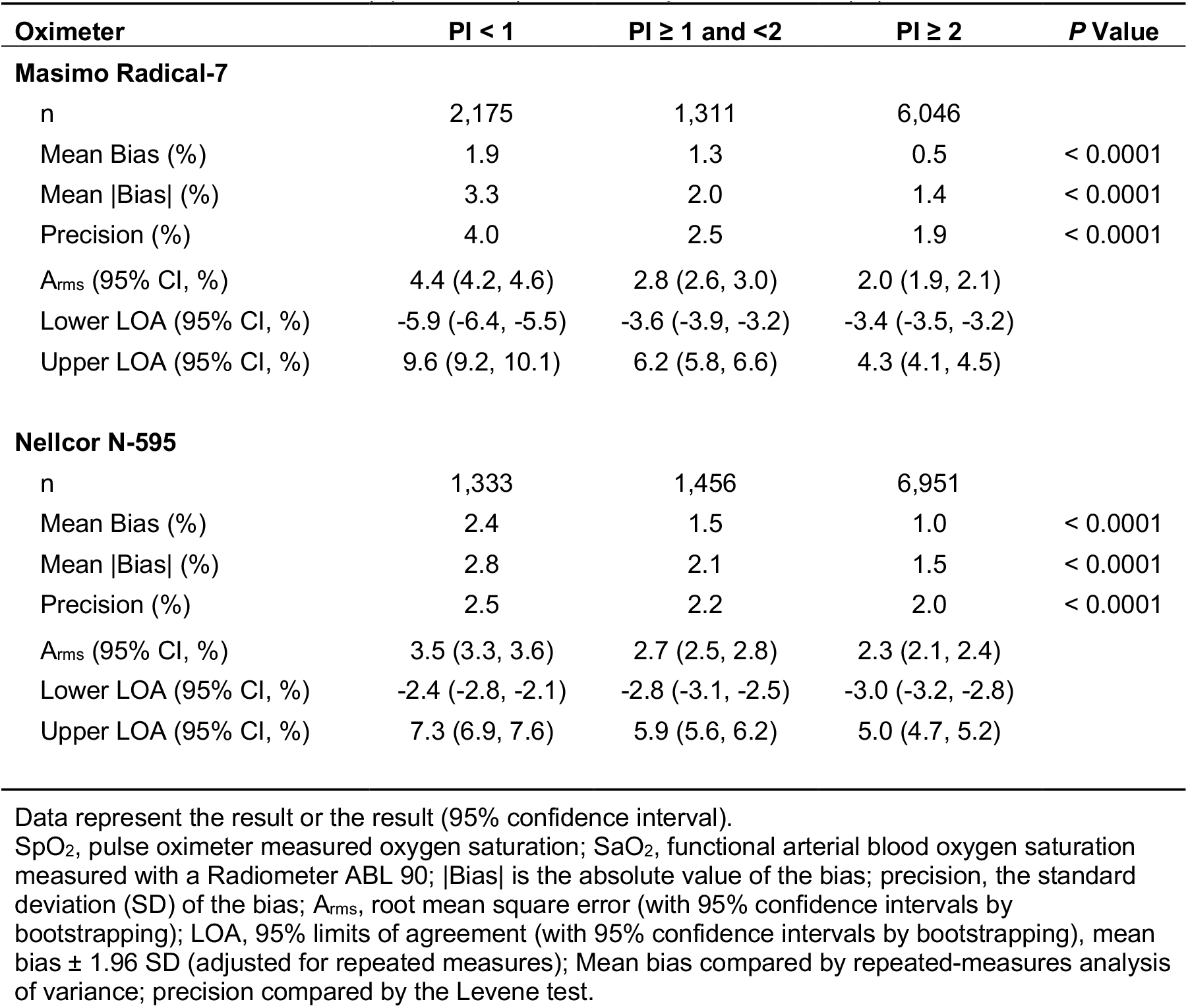
Pulse oximeter bias (SpO2 - SaO2) for different perfusion index (PI) levels.

**eTable 2.**
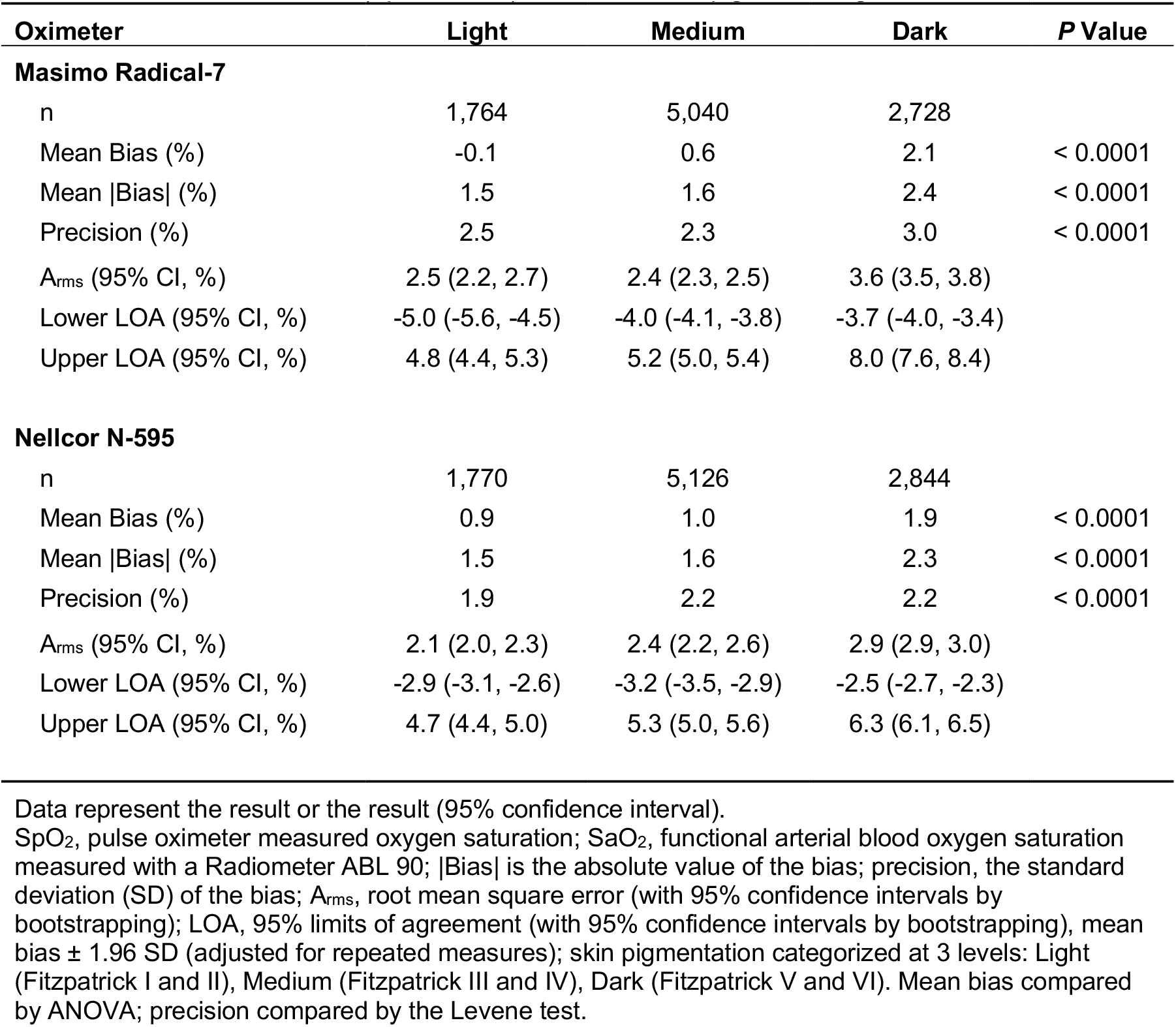
Pulse oximeter bias (SpO2 - SaO2) for different skin pigment categories

**eTable 3.**
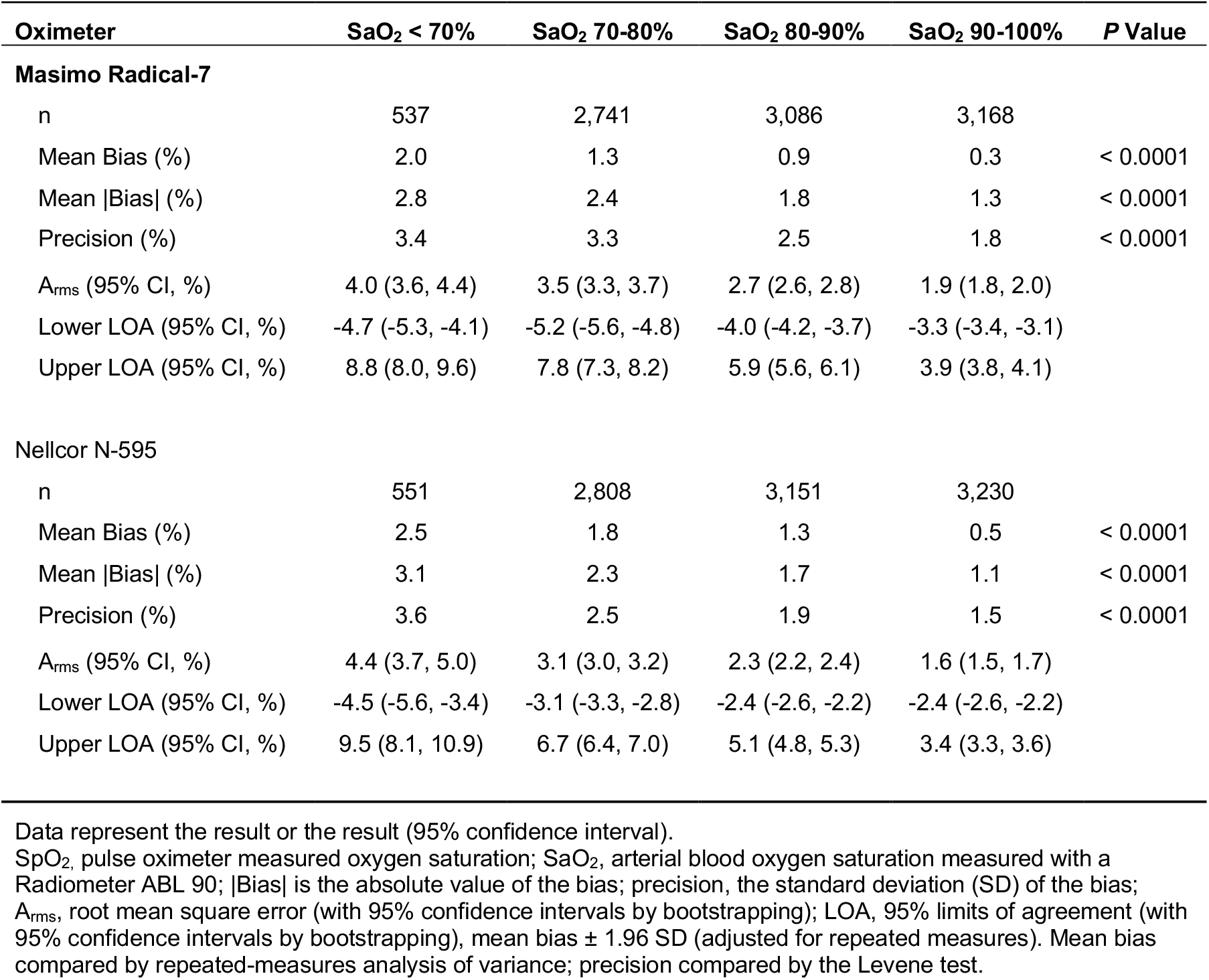
Pulse oximeter bias (SpO2 - SaO2) for different ranges of steady-state hypoxemia

**eTable 4.** Missed hypoxemia diagnosis, defined as SaO2 < 88%, with SpO2 at defined thresholds

| SpO<sub>2</sub> Threshold | ≥ 92% | ≥ 94% | ≥ 96% | 92-96% |
| --- | --- | --- | --- | --- |
| **Masimo Radical 7** |  |  |  |  |
| Light skin |  |  |  |  |
| PI < 1% | 1 (0.9%) | 0 (0.0%) | 0 (0.0%) | 1 (1.6%) |
| PI 1-2% | 1 (1.3%) | 0 (0.0%) | 0 (0.0%) | 1 (2.9%) |
| PI ≥ 2% | 1 (0.3%) | 1 (0.5%) | 1 (0.7%) | 0 (0.0%) |
| All Light | 3 (0.6%) | 1 (0.3%) | 1 (0.4%) | 2 (0.7%) |
| Medium skin |  |  |  |  |
| PI < 1% | 17 (4.2%) | 0 (0.0%) | 0 (0.0%) | 17 (8.5%) |
| PI 1-2% | 0 (0.0%) | 0 (0.0%) | 0 (0.0%) | 0 (0.0%) |
| PI ≥ 2% | 2 (0.2%) | 2 (0.3%) | 2 (0.5%) | 0 (0.0%) |
| All Medium | 19 (1.3%) | 2 (0.2%) | 2 (0.3%) | 17 (2.1%) |
| Dark skin |  |  |  |  |
| PI < 1% | 51 (19.1%) | 32 (14.3%) | 14 (8.7%) | 39 (30.2%) |
| PI 1-2% | 0 (0.0%) | 0 (0.0%) | 0 (0.0%) | 0 (0.0%) |
| PI ≥ 2% | 1 (0.2%) | 0 (0.0%) | 0 (0.0%) | 1 (0.4%) |
| All Dark | 52 (5.9%) | 32 (4.7%) | 14 (3.0%) | 40 (8.7%) |
| All | 74 (2.6%) | 35 (1.6%) | 17 (1.2%) | 59 (3.8%) |
| **Nellcor N-595** |  |  |  |  |
| Light skin |  |  |  |  |
| PI < 1% | 0 (0.0%) | 0 (0.0%) | 0 (0.0%) | 0 (0.0%) |
| PI 1-2% | 0 (0.0%) | 0 (0.0%) | 0 (0.0%) | 0 (0.0%) |
| PI ≥ 2% | 3 (0.7%) | 1 (0.4%) | 1 (0.5%) | 2 (0.8%) |
| All Light | 3 (0.5%) | 1 (0.2%) | 1 (0.3%) | 2 (0.6%) |
| Medium skin |  |  |  |  |
| PI < 1% | 9 (4.2%) | 2 (1.2%) | 0 (0.0%) | 9 (7.8%) |
| PI 1-2% | 1 (0.5%) | 0 (0.0%) | 0 (0.0%) | 1 (1.1%) |
| PI ≥ 2% | 8 (0.7%) | 8 (1.0%) | 8 (1.4%) | 0 (0.0%) |
| All Medium | 18 (1.2%) | 10 (0.9%) | 8 (1.0%) | 10 (1.2%) |
| Dark skin |  |  |  |  |
| PI < 1% | 7 (4.2%) | 3 (2.1%) | 0 (0.0%) | 7 (7.9%) |
| PI 1-2% | 1 (0.7%) | 0 (0.0%) | 0 (0.0%) | 1 (1.3%) |
| PI ≥ 2% | 4 (0.7%) | 0 (0.0%) | 0 (0.0%) | 4 (1.3%) |
| All Dark | 12 (1.4%) | 3 (0.5%) | 0 (0.0%) | 12 (2.5%) |
| All | 33 (1.1%) | 14 (0.6%) | 9 (0.6%) | 24 (1.4%) |

Missed hypoxemia is defined as the percentage of subjects in the specified SpO<sub>2</sub> range who have SaO<sub>2</sub> < 88%; SaO<sub>2</sub>, arterial blood oxygen saturation measured with a Radiometer ABL 90; SpO<sub>2</sub>, pulse oximeter measured oxygen saturation; Skin categories based on the Fitzpatrick scale: Light (I and II), Medium (III and IV), Dark (V and VI); PI was divided by 10 for the Nellcor N-595 to match scales

**eFigure 1.**
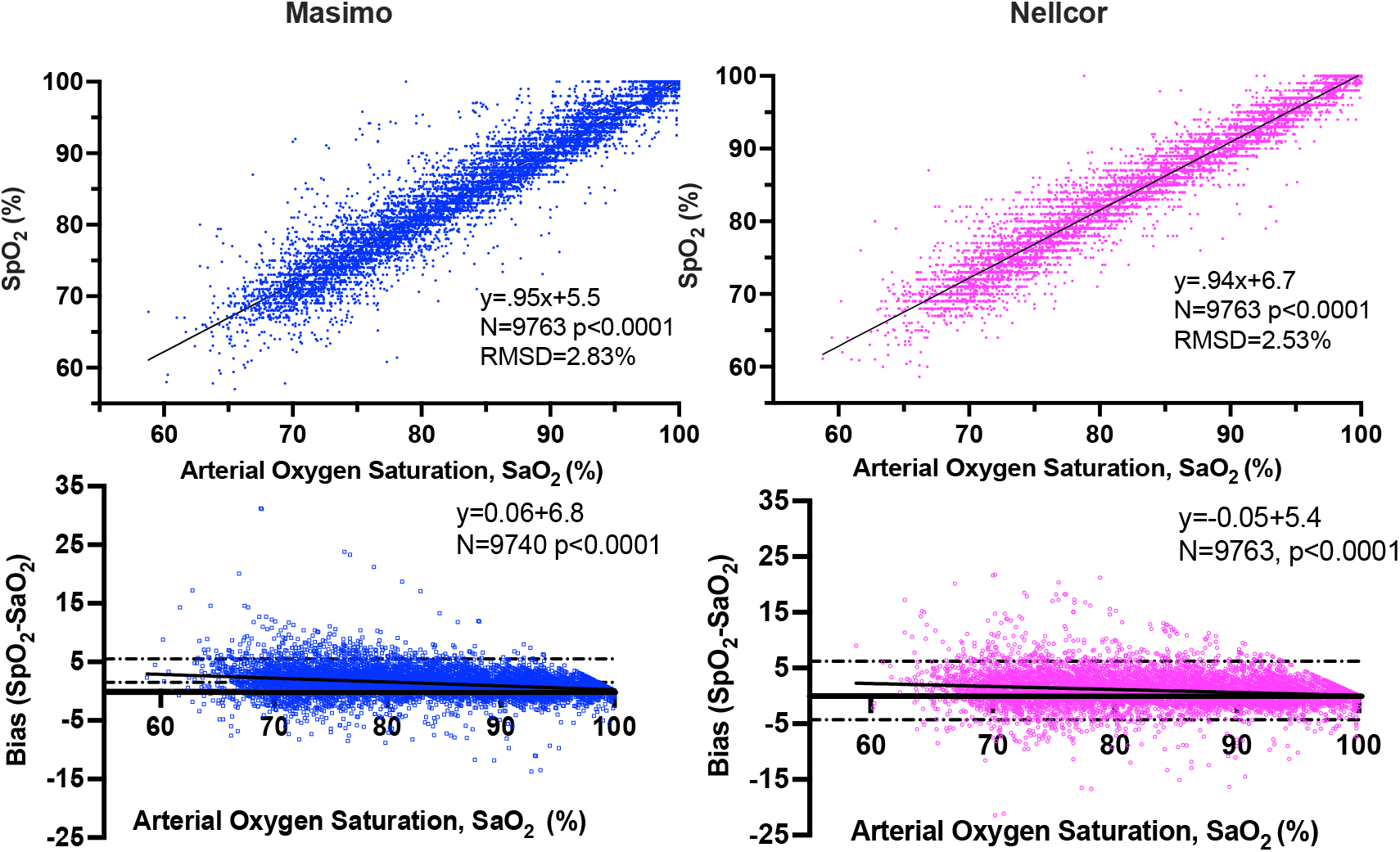
Scatterplot of pulse oximeter readings versus hemoximeter saturation values and pulse oximeter bias versus saturation. *Upper panels:* Point measurements of SaO_2_ (hemoximetry, arterial blood) and SpO_2_ readings from the Masimo Radical 7 and Nellcor N595 pulse oximeters during steady-state hypoxemia in healthy volunteers. The root mean-square deviation (precision) for the oximeters between 60-100% SaO_2_, including all subjects and conditions, was 2.94% for the Masimo and 2.67% for the Nellcor devices. *Lower Panels*: Modified Bland-Altman analysis of bias in 9763 pulse oximeter measurements. Lines indicate 95% confidence intervals (dashed) and least squares regression line.

## References

1. Jubran A, Tobin MJ. Reliability of pulse oximetry in titrating supplemental oxygen therapy in ventilator-dependent patients. Chest. 1990;97(6):1420–1425.

2. Severinghaus JW, Kelleher JF. Recent developments in pulse oximetry. Anesthesiology. 1992;76(6):1018–1038.

3. Batchelder PB, Raley DM. Maximizing the laboratory setting for testing devices and understanding statistical output in pulse oximetry. Anesth Analg. 2007;105(6 Suppl):S85–S94.

4. Bickler PE, Feiner JR, Lipnick MS, Batchelder P, MacLeod DB, Severinghaus JW. Effects of Acute, Profound Hypoxia on Healthy Humans: Implications for Safety of Tests Evaluating Pulse Oximetry or Tissue Oximetry Performance. Anesth Analg. 2017;124(1):146–153.

5. Bickler PE, Feiner JR, Severinghaus JW. Effects of skin pigmentation on pulse oximeter accuracy at low saturation. Anesthesiology. 2005;102(4):715–719.

6. Feiner JR, Severinghaus JW, Bickler PE. Dark skin decreases the accuracy of pulse oximeters at low oxygen saturation: the effects of oximeter probe type and gender. Anesth Analg. 2007;105(6 Suppl):S18-S23.

7. Pulse Oximeters-Premarket Notifications Submissions [510(k)s] Guidance for Industry and Food and Drug Administration Staff. In: FDA; 2007.

8. Okunlola OE, Lipnick MS, Batchelder PB, Bernstein M, Feiner JR, Bickler PE. Pulse Oximeter Performance, Racial Inequity, and the Work Ahead. Respir Care. 2022;67(2):252–257.

9. Andrist E, Nuppnau M, Barbaro RP, Valley TS, Sjoding MW. Association of Race With Pulse Oximetry Accuracy in Hospitalized Children. JAMA Netw Open. 2022;5(3):e224584.

10. Burnett GW, Stannard B, Wax DB, et al. Self-reported Race/Ethnicity and Intraoperative Occult Hypoxemia: A Retrospective Cohort Study. Anesthesiology. 2022;136(5):688–696.

11. Sjoding MW, Dickson RP, Iwashyna TJ, Gay SE, Valley TS. Racial Bias in Pulse Oximetry Measurement. N Engl J Med. 2020;383(25):2477–2478.

12. Valbuena VSM, Merchant RM, Hough CL. Racial and Ethnic Bias in Pulse Oximetry and Clinical Outcomes. JAMA Intern Med. 2022;182(7):699–700.

13. Pulse Oximeter Accuracy and Limitations: FDA Safety Communication. In:2021.

14. Warren E, Wyden R, Booker CA. Letter to the US FDA, 2021. https://www.warren.senate.gov/imo/media/doc/2020. Letter to FDA re Bias in Pulse Oximetry Measurements.pdf.

15. Henry NR, Hanson AC, Schulte PJ, et al. Disparities in Hypoxemia Detection by Pulse Oximetry Across Self-Identified Racial Groups and Associations With Clinical Outcomes. Crit Care Med. 2022;50(2):204–211.

16. Wong A-KI, Charpignon M, Kim H, et al. Analysis of Discrepancies Between Pulse Oximetry and Arterial Oxygen Saturation Measurements by Race and Ethnicity and Association With Organ Dysfunction and Mortality. JAMA Network Open. 2021;4(11):e2131674.

17. Louie A, Feiner JR, Bickler PE, Rhodes L, Bernstein M, Lucero J. Four Types of Pulse Oximeters Accurately Detect Hypoxia during Low Perfusion and Motion. Anesthesiology. 2018;128(3):520–530.

18. Pulse Oximeter Market by Product Global Forecast to 2027. 2022. https://www.marketsandmarkets.com/Market-Reports/pulse-oximeter-market-68168578.html. accessed 9/30/2022.

19. Premarket Notifications Submissions [510(k)s] Guidance for Industry and Food and Drug Administration Staff. FDA 2013.

20. Guideline for Evaluating and documenting SpO2 Accuracy in human Subjects. International Standards Organization; 2017.

21. Fitzpatrick TB. The Validity and Practicality of Sun-Reactive Skin Types I Through VI. Archives of Dermatology. 1988;124(6):869.

22. Bland JM AD. Statistical Methods for Assesing Agreement Between Two Methods of Clinical Measurements. The Lancet. 1986;327(8476):307–310.

23. Pulse Oximetry Errors in Hospitalized Patients Across Varying Skin Pigmentation (EquiOx). 2022. Clinical Trials.gov NCT 05554510.

24. Fawzy A, Wu TD, Wang K, et al. Racial and Ethnic Discrepancy in Pulse Oximetry and Delayed Identification of Treatment Eligibility Among Patients With COVID-19. JAMA Intern Med. 2022;182(7):730–738.

25. Falotico JM, Shinozaki K, Saeki K, Becker LB. Advances in the Approaches Using Peripheral Perfusion for Monitoring Hemodynamic Status. Front Med (Lausanne). 2020;7:614326.

26. UCSF Hypoxia Research Laboratory open source protocols. https://openoximetry.org/study-protocols/ accessed October 18, 2022.

